# Influencing Factors of Medical Doctors’ Intentions to Work in the Rural Health Facilities in the Eastern Cape Province, South Africa

**DOI:** 10.64898/2026.05.01.26352269

**Authors:** Simon Comley, Charity Masilela, Oladele Vincent Adeniyi

**Affiliations:** Department of Family Medicine and Rural Health, Faculty of Medicine & Health Sciences, Walter Sisulu University, South Africa; Lifestyle Diseases Research Focus Area, Faculty of Health Sciences, Northwest University, Mmabatho, South Africa

**Keywords:** Eastern Cape, Retention strategies, Resilience, Rural district hospitals, South Africa

## Abstract

**Background:** Understanding of context-specific retention strategies for doctors will guide targeted interventions and policy framework for strengthening the district health system in South Africa. Several strategies have been examined, some more impactful than others, with resilience playing a role in retention of staff, but data is lacking in the Eastern Cape Province, South Africa.

**Aim:** To assess factors influencing the retention and resilience of doctors at district hospitals in the Eastern Cape.

**Setting:** District hospitals in Amathole and Buffalo City health district municipalities in the Eastern Cape.

**Methods:** In this cross-sectional survey, participants rated retention strategies as well as a validated resilience scale (the CD-RISC 25).

**Results:** A total of 74 doctors were surveyed; mostly ≤34 years (66%), Black Africans (69%), and ≤5 years of professional experience (59%). The majority had worked in their current facilities for ≤5 years (76%). Significant proportion of young (78%), single (59%), and Grade 1 medical officers (86%) intend to leave their current facilities. Improving hospital accommodation was significantly associated with the intention to stay longer at the rural district hospitals. While not statistically significant, factors affecting professional development and growth scored higher while those related to financial remuneration scored lowest. There were no associations between resilience and intention to stay.

**Conclusion:** Early career doctors prioritise career growth and development, while more experienced doctors rated improved living condition as the main determinants of retention in the rural health facilities. Future studies should recruit representative sample of doctors from the various municipalities and across provinces in the country.

**Contribution:** Improving hospital accommodation and enhancing career growth and development may increase retention of doctors in the rural district hospitals.

## Introduction

Approximately 2000 new medical doctors complete their mandatory two-year internship training and proceed to community service in South Africa annually. These medical doctors are often young and relatively inexperienced, yet they form the backbone of healthcare service delivery in the rural district hospitals across the country. Even though these doctors are still early in their careers, they often face significant challenges during the community service year; heavy patient loads, insufficient resources, excessive work hours, isolation, and burnout. In addition, many patients present with advanced disease to the rural district hospitals, leading to challenges in medical management for junior doctors with limited clinical support.^1–7^ These challenges lead to significant pressure on junior doctors, spanning all aspects of their experiences, which could potentially impact their career decisions post-community service.^8, 9^

The district health system comprises primary healthcare clinics, community health centres and district hospitals. These facilities serve as the first point of contact for the majority of patients in the rural and semi-urban communities in South Africa. These facilities have a long history of staff shortages, inadequate funding for infrastructure and other important resources.^6, 10^ As such, the district health system depends largely on the annual distribution of the community service doctors, which is a mandatory requirement to become registered as an independent medical practitioner.^11^ While experienced doctors are needed to provide healthcare service as well as mentor the junior doctors in the district hospitals, there is a concern of high turnover of these senior doctors. This is often compounded by lack of funds to fill the vacated posts in the district hospitals.^8, 10, 11^ It is therefore pertinent to ascertain the reasons for the high turnover of doctors who have gained experience within these district hospitals, as well as examine possible retention strategies. By retaining doctors longer at the various district hospitals, more experienced clinicians will be available to transfer clinical skills to the junior doctors and consequently, improve the overall quality of healthcare service delivery.

Previous studies on the factors influencing retention of doctors have incorporated both focused questions on specific motivators for retention as well as the implications of burnout and resilience among medical doctors. Among others, financial incentives, support for career progression, hospital accommodation, support structures, management at the hospitals, resource availability, flexible working hours, improved specialist support, and enhanced social ties with the community are the main drivers of retention of doctors in the rural facilities.^1, 3, 9, 12^ Similarly, improving hospital infrastructure and enhancing hospital management are considered as stronger factors for retention than financial incentives in some studies.^3, 13–15^

In recent years, burnout among doctors has been linked to poor quality of healthcare service delivery, and thus, contributes to high turnover of staff from public health sector facilities. This has now become an important determinant of retention of doctors.^8, 16, 17^ Studies assessing burnout in South African doctors have shown a high correlation between burnout and adverse mental health of doctors, decreased ability to provide good patient care, and decreased likelihood to remain in the government sector.^8, 14, 16, 18^ On the other hand, resilience has emerged as a mitigating factor for burnout and has also been found to enhance the overall well-being of doctors working in rural facilities.^13, 16, 19, 20^

Despite several studies on retention strategies among doctors internationally as well as in South Africa,^8, 13, 14, 16, 17, 21, 22^ there is a paucity of research in the Eastern Cape, a resource-poor and understudied province. The Eastern Cape province is the fourth most populous province in South Africa with 58% of its population living in the rural communities. The province has the highest number of households dependent on social grants, poor health service, high levels of litigation-related debt, and decreased confidence in the health system by communities.^23–25^ Specifically, the Amathole health district has a higher number of people living in the rural communities than the Buffalo City Municipality (80% and 41.6%, respectively) with both municipalities serving large numbers of people sparsely distributed.^26^ In order to strengthen the district health system across the Eastern Cape province, a focused, context-specific study addressing the determinants of retention of more experienced doctors at this level of care is critical.

With better knowledge of these factors, policy-makers could be informed more comprehensively of potential interventions to ensure better staffing at the facilities, improved health and capabilities of the doctors, and an enhanced provision of quality primary health care to these vulnerable patients. This study determines the influencing factors of retention in the rural district hospitals in the Amathole and Buffalo City Municipalities of the Eastern Cape. Additionally, the study assesses the resilience levels of doctors working in the region and the association between resilience levels and intention to continue working in the rural health facilities.

## Methods

### Study design and setting

This descriptive cross-sectional study was conducted between 01 October 2024 to 31 March 2025 across the district hospitals in the Amathole (n=12) and Buffalo City (n=2) Municipalities in the central region of the Eastern Cape. The two district hospitals in the Buffalo City Metropolitan Municipality are located in the semi-urban areas in comparison to the 12 district hospitals located in the predominantly rural communities across the entire Amathole district. These hospitals are manned by medical doctors with varying years of experience; from community service doctors to those who have worked for over 20 years. The majority of district hospitals in the region have accommodation for the doctors as part of their retention strategies and to facilitate quicker access for emergency services. These doctors are supported by the district clinical specialist team: Family physicians, Paediatricians, Obstetrics & Gynaecology specialists and Psychiatrists. These higher cadres of doctors provide clinical governance and expert guidance in patient care at the district hospitals.

### Ethical Consideration

The Walter Sisulu University Research Ethics Committee granted protocol approval for the study (Approval number: 164/2024). Permission for the implementation of this study was obtained from the Eastern Cape Department of Health and the clinical governance of each hospital. No participants’ identifiers were obtained during the study. Each participant completed written informed consent either in hard or soft copy (electronic) demonstrating willingness to participate in the study. The study did not pose any threat to patient care, however, there was a chance of influencing the career choices of doctors surveyed by introspection of work difficulties and experiences. A toll-free support line contact number was provided in the participant information sheet for assistance for any doctor who may be affected during the study.

### Study population and sample size estimation

All 118 medical doctors registered on the PERSAL system of the Eastern Cape Department of Health for the 14 district hospitals were eligible for the study. A sample size of 91 was initially estimated, using Slovin’s formula;

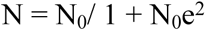

(where *N_0_ is the total population size, e is the margin of error and considered as 5%)*.

However, in anticipation of non-response and/or incomplete responses to important outcome measures from the participants, the entire team of doctors were sampled using census sampling technique.

### Data collection

In order to ensure maximal participation of the doctors, the clinical managers of the hospitals were contacted and facilitated access to their team. Participant’s information sheet detailing the purpose and process of the study was distributed by the clinical managers to their respective team of doctors prior to recruitment. After obtaining written informed consent, each participant completed a self-administered questionnaire within 25 minutes between October 2024 to March in 2025.

The questionnaire was distributed via a Google Form (electronic copy) to encourage completion remotely by the participants at their convenience. Numerous follow-ups with the doctors were made electronically (via email reminders) as well as scheduled visits to the various hospitals. Allowance was made for doctors who were on annual leave by extending the period of data collection. Responses from the electronic forms were captured into a secured, password-protected computer.

### Measures

The questionnaire comprised socio-demographic characteristics (age, sex, race, place of birth, relationship status, whether they have children or not and professional category), information about rural versus urban upbringing, medical training, and internship. These variables have been shown to predict retention of doctors in specific geographic areas.^27^ Additional information on retention strategies employed by the doctors were elicited based on previous reports from the literature.^9, 12, 13, 21^ Some of these factors include: increased pay in rural areas, better teamwork, opportunities for professional development, improving hospital accommodation, and increasing staffing numbers.^9, 13, 21^

Similarly, a previously validated Connor Davidson Resilience scale (CD-RISC), which comprises the 25-item measure for assessing doctors’ resilience was included. Briefly, the scale requires the respondent to rate a variety of questions regarding how they react to certain events or cope with difficulties, these are synthesized into a total resilience score as well as scores within groups of resilience domains. These domains can then be grouped into a variety of sub-themes as explored in different literature to provide insight into the phenomenon of interest.^28^.

The CD-RISC 25 Resilience Scale rating has been validated for use in South Africa with a Cronbach alpha score of 0.89.^20, 29^ Nonetheless, the questionnaire was piloted with 10 doctors in the department of family medicine at the regional hospital in the central region of the Eastern Cape to assess user-friendliness and ease of completion. The instrument was judged to be suitable and user-friendly. The results from the pilot were not included in the final analysis.

## Data analysis

All analyses were conducted using IBM SPSS Statistics for Windows, Version 27.0 ((IBM Corp., Armonk, New York, USA). Data were inspected and cleaned prior to analysis, and the distribution of continuous variables was assessed using the Shapiro–Wilk test. For consistency, all continuous variables, including motivation and resilience dimension scores, are presented as means ± standard deviations (SD). Relationships between categorical variables (e.g., socio-demographics and intention to stay; socio-demographics and duration of service at the current facility) were examined using chi-square tests. The individual hospital-related factors were grouped into five motivation dimensions: Hospital Infrastructure & Resources, Professional Development & Growth, Financial & Job Security, Interpersonal Work Relations, and Management & System Strengthening. The motivation dimension scores were analysed in relation to intention to stay (Yes/No) by using Mann–Whitney U tests. Resilience dimension scores were categorised as; Tenacity and Competence, Trust, Acceptance of Change and Secure Relationships, Control and Spirituality in accordance to previous study [30], and were compared across the sociodemographic variables using independent t-tests or one-way ANOVA (for three groups)^30^. Tukey’s Honest Significant Difference (HSD) post-hoc test was applied following significant one-way ANOVA to identify group differences while controlling for Type I error. All p-values < 0.05 were considered statistically significant..

## Results

### Demographic and Professional Characteristics of the Study Cohort

A total of 74 medical doctors participated in the study, including nine from the Buffalo City Metropolitan Municipality and sixty-five from the Amathole district. The cohort comprised 43.2% males (n = 32) and 54.05% females (n = 40). The majority of participants were Black (68.92%), aged ≤34 years (66.2%), married (52.7%), and had ≤5 years of professional experience as medical doctors (59.5%). In addition, 75.7% (n = 56) had been serving at their current facility for ≤5 years, and 67.6% (n = 50) were in the Grade 1 professional category. The chi-square analysis revealed significant associations between socio-demographic characteristics; age (p < 0.001), professional category (p < 0.001), community service location (p = 0.024), years of experience as a medical doctor (p < 0.001), and duration of service at the current facility (Table 1).

**Table 1:** Demographic characteristics of participants.

| Variable | Total (n; %) | Duration of Service at the Current Facility |  | p-values |
| --- | --- | --- | --- | --- |
|  |  | ≤5 years (n; %) | >5 years (n; %) |  |
| **All** | 74 (100) | 56 (75.7) | 18 (24.3) |  |
| **Age** |  |  |  | **<0.001** |
| ≤34 years | 49 (66.2) | 44 (78.6) | 5 (27.8) |  |
| >34 years | 25 (33.8) | 12 (21.4) | 13 (72.2) |  |
| Mean±SD | 34.09±7.87 | 31.41±4.27 | 42.06±10.77 |  |
| **Sex** |  |  |  | 1.000 |
| Male | 32 (43.2) | 24 (42.9) | 8 (44.4) |  |
| Female | 40 (54.1) | 30 (53.6) | 10 (55.6) |  |
| **Race** |  |  |  | 0.411 |
| Black | 51 (68.9) | 40 (71.4) | 11 (61.1) |  |
| White | 23 (31.1) | 16 (28.6) | 7 (38.9) |  |
| **Place of Birth** |  |  |  | 0.423 |
| Rural | 39 (52.7) | 31 (55.4) | 8 (44.4) |  |
| Urban | 35 (47.3) | 25 (44.6) | 10 (55.6) |  |
| **Relationship Status** |  |  |  | 0.610 |
| Single | 34 (45.9) | 27 (48.2) | 7 (38.9) |  |
| Married | 39 (52.7) | 29 (51.8) | 10 (55.6) |  |
| **Already have children** |  |  |  | 0.075 |
| Yes | 34 (45.9) | 29 (51.8) | 5 (27.8) |  |
| No | 40 (54.1) | 27 (48.2) | 13 (72.2) |  |
| **Professional category** |  |  |  | <0.001 |
| Grade 1 | 50 (67.6) | 47 (83.9) | 3 (16.7) |  |
| Grade 2 | 10 (13.5) | 3 (5.4) | 7 (38.9) |  |
| Grade 3 | 14 (18.92) | 6 (10.7) | 8 (44.4) |  |
| **Current Hospital Location** |  |  |  | 0.133 |
| BMC | 09 (12.1) | 5 (8.9) | 4 (22.2) |  |
| Amathole | 65 (87.8) | 51 (91.1) | 14 (77.8) |  |
| **Community Service Location** |  |  |  | 0.024 |
| Rural | 54 (72.9) | 45 (80.4) | 9 (50.0) |  |
| urban | 19 (25.7) | 11 (19.6) | 8 (44.4) |  |
| **Internship Location** |  |  |  | 0.457 |
| Rural | 13 (17.6) | 11 (19.6) | 2 (11.1) |  |
| Urban | 60 (81.1) | 45 (80.4) | 15 (83.3) |  |
| **Experience as a Doctor (Years)** |  |  |  | <0.001 |
| ≤ 5 years | 44 (59.5) | 44 (78.6) | 0 (0.00) |  |
| >5 years | 30 (40.5) | 12 (21.4) | 18 (100) |  |

Of the 74 participants, 50% (n = 37) indicated an intention to remain at their current facility. Chi-square analysis revealed that age (p = 0.027), relationship status (p = 0.025), level of qualification (p = 0.002), and years of service at the current facility (p = 0.030) were significantly associated with the intention to stay. No significant associations were observed for other variables (Table 2).

**Table 2:** Intention of doctors to continue working in the rural facilities.

| Variable | Total<br>(n; %) | Intention to Stay |  | p-values |
| --- | --- | --- | --- | --- |
|  |  | Yes (n; %) | No (n; %) |  |
| **All** | 74 (100) | 37 (50.0) | 37 (50.0) |  |
| **Age** |  |  |  | **0.027** |
| ≤34 years | 49 (66.2) | 20 (54.1) | 29 (78.4) |  |
| >34 years | 25 (33.8) | 17 (45.9) | 8 (21.6) |  |
| **Sex** |  |  |  | 0.225 |
| Male | 32 (43.2) | 19 (51.4) | 13 (35.1) |  |
| Female | 40 (54.1) | 18 (48.7) | 22 (59.5) |  |
| **Race** |  |  |  | 0.451 |
| Black | 51 (68.9) | 24 (64.9) | 27 (72.9) |  |
| White | 23 (31.08) | 13 (35.1) | 10 (27.0) |  |
| **Place of Birth** |  |  |  | 0.224 |
| Rural | 39 (52.7) | 22 (59.5) | 17 (45.9) |  |
| Urban | 35 (47.30) | 15 (40.5) | 20 (54.1) |  |
| **Relationship Status** |  |  |  | **0.025** |
| Single | 34 (45.9) | 12 (32.4) | 22 (59.5) |  |
| Married | 39 (52.7) | 24 (64.9) | 15 (40.5) |  |
| **Already have children** |  |  |  | 0.351 |
| Yes | 34 (45.9) | 15 (40.5) | 19 (51.4) |  |
| No | 40 (54.1) | 22 (59.5) | 18 (48.7) |  |
| **Level of Qualification** |  |  |  | **0.002** |
| Grade 1 | 50 (67.6) | 18 (48.7) | 32 (86.5) |  |
| Grade 2 | 10 (13.5) | 7 (18.9) | 3 (8.1) |  |
| Grade 3 | 14 (18.9) | 12 (32.4) | 2 (5.4) |  |
| **Current Hospital Location** |  |  |  | 0.772 |
| BMC | 9 (12.2) | 5 (13.5) | 4 (10.8) |  |
| Amathole | 65 (87.8) | 32 (86.5) | 33 (89.2) |  |
| **Community Service Location** |  |  |  | 0.161 |
| Rural | 54 (72.9) | 24 (64.9) | 30 (81.1) |  |
| Urban | 19 (25.7) | 12 (32.4) | 7 (18.9) |  |
| **Internship Location** |  |  |  | 0.113 |
| Rural | 13 (17.6) | 9 (24.3) | 4 (10.8) |  |
| Urban | 60 (81.1) | 27 (72.9) | 33 (89.2) |  |
| **Experience as a Doctor (Years)** |  |  |  | 0.058 |
| ≤ 5 years | 44 (59.5) | 18 (48.7) | 26 (70.3) |  |
| >5 years | 30 (40.5) | 19 (51.4) | 11 (29.7) |  |
| **Number of Years at the Current Facilities** |  |  |  | **0.030** |
| ≤ 5 years | 56 (75.7) | 24 (64.9) | 32 (86.5) |  |
| >5 years | 18 (24.3) | 13 (35.1) | 5 (13.5) |  |

The mean scores for the Trust dimension ranged from 2.69 ± 0.54 to 2.97 ± 0.61, while Acceptance of Change and Secure Relationships scores ranged from 2.80 ± 0.59 to 3.47 ± 0.80 across participant subgroups. Independent t-test revealed significant differences in Acceptance of Change and Secure Relationships by age (p = 0.044) and race (p = 0.044), with participants ≤34 years (3.03±0.68) and White (3.47±.59) scoring higher in this dimension. No other demographic or professional variables showed significant differences in either the Trust or Acceptance of Change & Secure Relationships dimensions (Table 3).

**Table 3:**
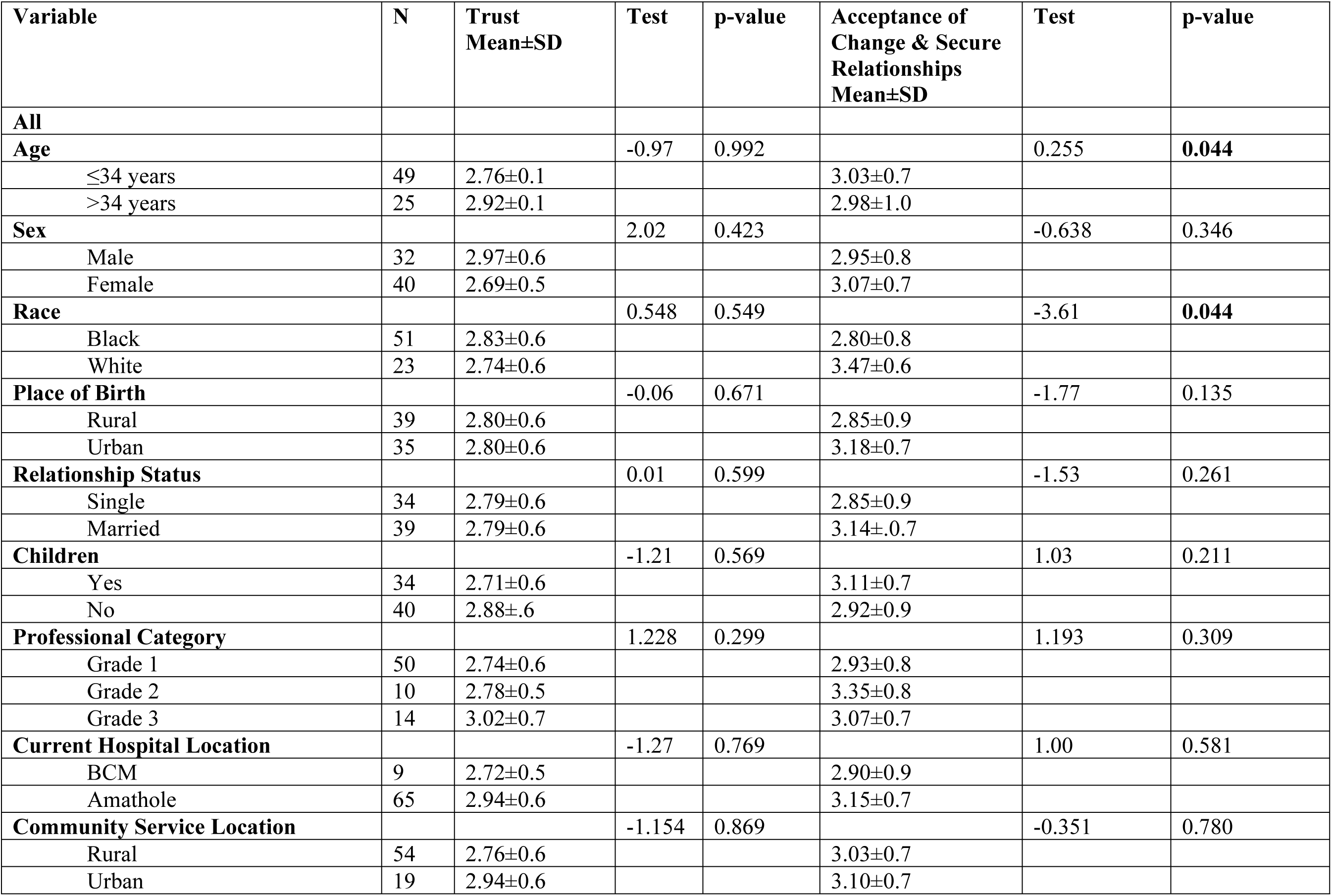
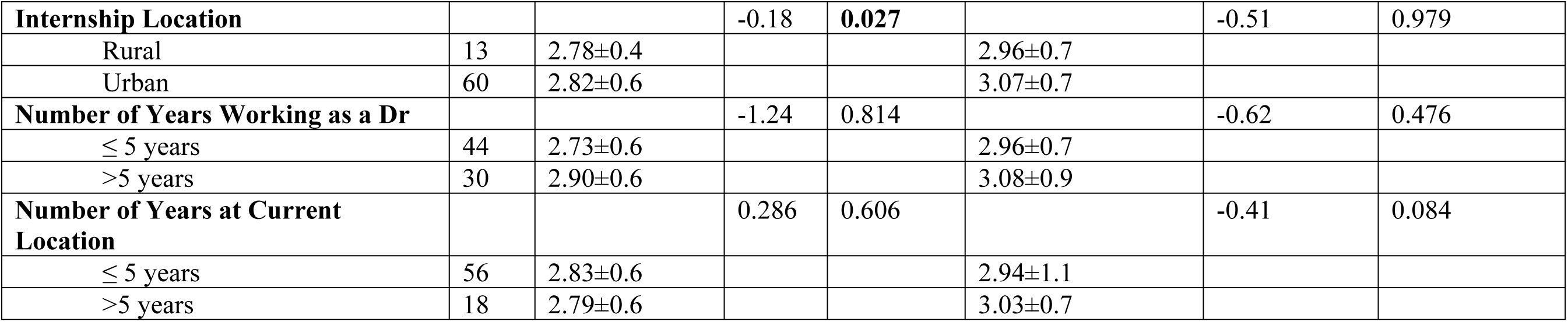
Resilience dimensions [Independent t-test and ANOVA (for 3 groups)]

The mean scores for the Control and Spirituality dimension ranged from 2.52 ± 0.60 to 3.07 ± 0.76, while scores for Tenacity and Competence ranged from 2.61 ± 0.41 to 3.80 ± 0.42 across the various subgroups. For Control and Spirituality, the independent t-test revealed a significant difference in scores by place of birth (rural vs. urban; p = 0.026). Similarly, the one-way ANOVA indicated a significant effect of professional category (Grade 1–3; p = 0.043). However, Tukey’s post-hoc analysis did not identify any significant pairwise differences between professional category. Overall, participants born in urban areas (2.70± 0.75) and those with Grade 2 level (3.07±0.76) scored higher on this dimension compared to their counterparts. Additionally, significant differences in Tenacity and Competence scores were observed by race (p = 0.036), relationship status (single vs. married; p = 0.014), and parental status (having children; p = 0.003). Specifically, black participants (2.92±0.64), married participants (2.92±0.40), and those without children (3.02±0.43) had higher mean scores on this dimension. No significant differences were found for age, sex, years of experience as a medical doctor, or current facility location (Table 4).

**Table 4:** Resilience dimensions [Independent t-test and ANOVA (for 3 groups)]

| Variable | N | Control & Spirituality<br>Mean±SD | Test | p-value | Tenacity & Competence<br>Mean±SD | Test | p-value |
| --- | --- | --- | --- | --- | --- | --- | --- |
| **All** |  |  |  |  |  |  |  |
| **Age** |  |  | -1.14 | 0.572 |  | -0.88 | 0.278 |
| ≤34 years | 49 | 2.56±0.6 |  |  | 2.87±0.6 |  |  |
| >34 years | 25 | 2.75±0.7 |  |  | 2.99±0.5 |  |  |
| **Sex** |  |  | -0.42 | 0.688 |  | 1.14 | 0.618 |
| Male | 32 | 2.60±0.7 |  |  | 3.01±0.6 |  |  |
| Female | 40 | 2.66±0.6 |  |  | 2.86±0.5 |  |  |
| **Race** |  |  | -0.40 | 0.652 |  | 0.27 | **0.036** |
| Black | 51 | 2.60±0.6 |  |  | 2.92±0.6 |  |  |
| White | 23 | 2.67±0.7 |  |  | 2.89±0.4 |  |  |
| **Place of Birth** |  |  | -0.89 | **0.026** |  | 0.50 | 0.577 |
| Rural | 39 | 2.56±0.6 |  |  | 2.94±0.6 |  |  |
| Urban | 35 | 2.70±0.7 |  |  | 2.89±0.5 |  |  |
| **Relationship Status** |  |  | 0.28 | 0.557 |  | -0.25 | **0.014** |
| Single | 34 | 2.63±0.6 |  |  | 2.88±0.7 |  |  |
| Married | 39 | 2.59±0.7 |  |  | 2.92±0.4 |  |  |
| **Children** |  |  |  |  |  |  | **0.003** |
| Yes | 34 | 2.66±0.6 | 0.49 | 0.707 | 2.78±0.7 | -1.82 |  |
| No | 40 | 2.59±0.7 |  |  | 3.02±0.4 |  |  |
| **Professional Category** |  |  | 3.28 | **0.043** |  | 1.38 | 0.256 |
| Grade 1 | 50 | 2.52±0.6 |  |  | 2.91±0.6 |  |  |
| Grade 2 | 10 | 3.07±0.8 |  |  | 2.68±0.7 |  |  |
| Grade 3 | 14 | 2.69±0.7 |  |  | 3.80±0.4 |  |  |
| **Current Hospital Location** |  |  | 0.22 | 0.505 |  | 1.20 | 0.123 |
| BCM | 9 | 2.59±0.7 |  |  | 2.61±0.6 |  |  |
| Amathole | 65 | 2.52±0.7 |  |  | 3.00±0.7 |  |  |
| **Community Service Location** |  |  | -1.18 | 0.167 |  | -0.81 | 0.196 |
| Rural | 54 | 2.58±0.6 |  |  | 2.87±0.5 |  |  |
| Urban | 19 | 2.78±0.7 |  |  | 3.00±0.7 |  |  |
| **Internship Location** |  |  | 1.63 | 0.444 |  | -1.42 | 0.880 |
| Rural | 13 | 2.90±0.6 |  |  | 2.70±0.6 |  |  |
| Urban | 60 | 2.57±0.7 |  |  | 2.95±0.6 |  |  |
| **Number of Years as a doctor** |  |  | -1.60 | 0.298 |  | -0.66 | 0.319 |
| ≤ 5 years | 44 | 2.52±0.6 |  |  | 2.87±0.6 |  |  |
| >5 years | 30 | 2.77±0.7 |  |  | 2.97±0.5 |  |  |
| **Number of Years at Current Location** |  |  | 0.49 | 0.757 |  | 0.60 | 0.894 |
| ≤ 5 years | 56 | 2.69±0.7 |  |  | 2.98±0.6 |  |  |
| >5 years | 18 | 2.60±0.6 |  |  | 2.89±0.6 |  |  |

Analysis of motivation dimensions in relation to participants’ intention to stay revealed no significant differences between those who intended to stay and those who did not. Mean scores were consistently high across all dimensions: Hospital Infrastructure and Resources (Yes: 4.07 ± 0.84), Professional Development and Growth (4.54 ± 0.73), Financial and Job Security (3.85 ± 0.58), Interpersonal Work Relations (4.17 ± 1.00), as well as Management and System Strengthening (4.33 ± 0.78) (Table 5).

**Table 5:** Retention strategy categories (Independent sample Mann-Whitney U Test)

| Motivation Dimension | Intention to Stay |  | Test | p-value |
| --- | --- | --- | --- | --- |
| | Yes (Mean±SD) | No (Mean±SD) | | |
| Hospital Infrastructure & Resources | 4.07±0.84 | 4.14±0.68 | 687.00 | 0.978 |
| Professional Development & Growth | 4.54±0.73 | 4.60±0.71 | 762.50 | 0.359 |
| Financial & Job Security | 3.85±0.58 | 3.71±0.42 | 599.00 | 0.341 |
| Interpersonal Work Relations | 4.17±1.00 | 3.95±.89 | 544.50 | 0.125 |
| Management & System Strengthening | 4.33±0.78 | 4.34±0.79 | 713.50 | 0.747 |

Analysis of the retention strategy categories demonstrated the highest rating being Professional Development and Growth (4.57±0.72), followed by Management and System Strengthening (4.33±0.78), Hospital Infrastructure and Resources (4.11±0.76), Interpersonal Work Relations (4.06±0.95) with the lowest being Financial & Job Security (3.78±0.51) (Table 6).

**Table 6:** Retention strategy groups Mean Score (n=74)

| Retention strategy groups | N | Minimum | Maximum | Mean±SD | Normality Assumption (p-Values) |
| --- | --- | --- | --- | --- | --- |
| Hospital Infrastructure & Resources | 74 | 1.00 | 5.00 | 4.11±0.76 | <0.001 |
| Professional Development & Growth | 74 | 1.00 | 5.00 | 4.57±0.72 | <0.001 |
| Financial & Job Security | 74 | 2.33 | 5.00 | 3.78±0.51 | 0.004 |
| Interpersonal Work Relations | 74 | 1.00 | 5.00 | 4.06±0.95 | <0.001 |
| Management & System Strengthening | 75 | 1.00 | 5.00 | 4.33±0.78 | <0.001 |

Chi-square analysis showed no significant association between intention to stay and the hospital factors/variables (p > 0.05). However, a significant association was observed between duration of service in the current facility and improvement in hospital accommodation (p = 0.026) (Supplementary Table 1)

## Discussion

The health system of the Eastern Cape has many challenges: a widely dispersed and impoverished population depends on the district hospitals for most of their care. These health facilities experience many problems; staff shortage, drug stock-out, poor management, and inadequate support for junior doctors.^4, 6, 10, 31^ The annual rotation of doctors away from these facilities at the end of their community service year to other areas leads to a more challenging environment for ongoing healthcare teams and patient support. It is vital to understand the factors that assist with retention of doctors in these facilities thus laying the foundation for human resources for health discourse with policymakers to undertake appropriate actions to enable better continuity of skills and care at these facilities. Previous research has suggested that the best time to implement strategies for retention is in the first year of practice – predicting longer retention – this further emphasises the importance of exploring retention strategies among junior doctors in particular.^13^

### Demographics and intention to stay

Of interest in the present study is that young and early career medical doctors indicated their intention to leave their rural hospital posts. These cadre of doctors constitute half of the respondents in the study. The concern here is that the doctors who have the potential to develop themselves and engage in longitudinal upliftment of the hospital, leave before their worth can be realised along with the benefits they can bring. If a doctor grows within a hospital, they could also grow aspects of the hospital in terms of sharing skills with colleagues. Indeed, in research assessing retention of Community Service doctors in district hospitals within the KwaZulu-Natal, Eastern Cape, and Limpopo provinces the authors report that the annual exodus of Community Service doctors leads to a loss of the skills and experience gained during that year. These doctors are valuable assets worth retaining in these hospitals and their loss would negatively impact patient care and service delivery.^32^

While it is expected that the rural upbringing of doctors or prior exposure to the rural facilities during medical internship or community service would shape the intentions or desire of doctors to work in the rural areas, these results suggest otherwise with no significant association between intention to stay, and rural upbringing and rural internship allocation. This finding corroborates previous reports within South Africa by Mash^22^ in 2022 and Baytopp^27^ in 2025. However, this differs from earlier reports from 2009 by Ross & Reid^32^ from the KwaZulu Natal province which demonstrated that Community Service doctors who remained in rural district hospitals had rural upbringing.

It should be noted that research in other countries has recommended targeted admission policies aimed at enrolling students from rural background into medical schools with a view to deploying them to serve the rural communities once their studies are completed.^13^ These strategies have yielded good outcomes in other regions, such as: Australia, Europe, India, and North America.^9,13, 27^ The WHO also recommends targeted admission strategies as well as exposing medical students to rural contexts during their training to enhance retention of doctors in the rural areas.^13, 33^

It is surprising that half of all the respondents intend to remain in their current rural district hospital posts. This finding represents a higher proportion of medical doctors desiring to continue working in the rural hospitals compared to previous reports from South Africa; ranging between 8% and 23%. ^27, 34^ This may reflect current landscape of decreased availability of medical doctors’ posts elsewhere thus, dissuading some doctors from leaving their current facility. Among the young doctors and those early in their career in the present study, 22% indicated their intention to continue working in the rural district hospitals. This finding is roughly similar to previous reports analysing retention of junior doctors in South Africa where approximately 15% of community service doctors and approximately 23% of Grade 1 MOs reported an intention to remain in the rural areas.^27, 35, 36^

### Primary retention strategies Improving hospital accommodation

The only strategy showing a significant positive effect on intention to stay in the rural hospital was improving hospital accommodation as it applied to doctors who had been working at that facility for more than five years. This suggests that the longer a doctor stays at a facility, the greater importance they place on their accommodation. This finding corroborates findings from other studies on the association between length of stay and accommodation. A study conducted in a different province in South Africa demonstrated that a greater proportion of rural health professionals lived in hospital accommodation compared to their urban counterparts.^37^ However, a substantial proportion of the doctors (59%) who live in hospital accommodation reported dissatisfaction; they perceived the living and working conditions to be poor.^37^

This is important when considering length of stay because a study assessing predicted length of service in the district health system demonstrated an increasing likelihood of retention as doctors grew through their post – only 5% of those who had practiced for less than one year planned to stay while 41% of those who had stayed for more than four years planned to stay in their post.^9^ This same study found that doctors not being able to live with their spouse and not having a strong local support system were significantly associated with desire to leave their district hospital post.^9^ Another study in South Africa specifically mentioned that many community service doctors intended to leave their facilities purely because of poor hospital accommodation.^38^ As such, accommodation conducive for the doctors and their family could be an incentive for doctors to consider staying longer in the rural facilities. This recommendation has also been echoed in several studies globally, specifically perceptions of accommodation status are linked to deprivation rate of an area, lack of amenities, deficiencies in infrastructure and poor contact with family negatively impacted retention of doctors in the rural facilities.^1, 35, 39^

### Professional development and growth

Of interest is the high rating of professional development and growth score among the respondents. Though, this dimension did not show any significant association with the intention to stay in the rural hospitals in the present study. This finding is hardly surprising, given that the majority of the respondents were young and early career practitioners. The high rating of professional development and growth reflects the ambition of the young doctors, who typically would pursue postgraduate training in the tertiary hospitals. This finding corroborates previous studies that have linked career growth and development as reasons for doctors leaving their post.^1, 13, 27, 40^ Findings from previous research in South Africa showed that doctors with the desire to specialise in certain fields are more likely to leave the district health system in the Western Cape.^6^ Similarly, a study from the KwaZulu-Natal Province in South Africa showed that only few doctors were able to study towards further qualifications in the rural hospitals, but interestingly, this did not affect their desire to remain at the rural hospital. ^37^

### Financial and job security

Financial and job security was rated the lowest retention strategy by the respondents in this study, though, no significant association was established with the intention to stay in the rural facilities. This result should be treated with caution, given the small sample size of the respondents. It should be noted that there are mixed reports in the literature on the relationship between the intention to stay in the rural facilities and financial motivation. While several studies have reported that aspects pertaining to financial security were rated the lowest among the factors influencing intention to stay in rural areas, other studies outside of South Africa found that financial incentives did play a significant role in retention of doctors.^9, 27, 38^ In fact, a systematic review concluded that financial incentives for retention of doctors in rural communities deserve the attention of policymakers.^1^ Perhaps, the rural allowance paid to doctors working in the rural facilities across South Africa may have influenced the rating of this particular dimension and as such, financial incentives were no longer considered an attractive retention strategy.

### Resilience dimensions

Analysis of the various resilience dimensions in this study revealed significant findings in each of the dimensions. Those who had been interns in urban areas displayed significantly higher scores in the Trust dimension. Those who were 34 years or younger and those who were White showed significantly higher scores in the dimension of Acceptance of Change and Secure Relationships. Those who were born in urban areas and those who were in Grade 2 MO positions showed significantly higher scores in the dimension of Control and Spirituality. Those doctors who were Black, married, and had children all showed significantly higher scores in the dimension of Tenacity and Competence. These resilience dimensions provide insight into the participants, however none of these dimensions demonstrated a significant relationship with intention to stay. A study in South Africa showed that, second to working conditions, mental health and resilience played a significant role in increasing the likelihood of a doctor to work in the rural areas.^27^ Resilience levels play a role in preventing burnout and thus, assists with a higher likelihood of retention of doctors in challenging contexts including urban but more especially rural areas.^16, 17,20^ Prior studies have demonstrated that higher resilience scores predict higher likelihood of retention, lower levels of stress and burnout, and improved workplace functioning with resultant positive impact on improving patient care.^41, 42^

### Study limitations

This study was conducted in two of the total of 8 health districts in the Eastern Cape with a small sample of medical doctors. As such, findings should be interpreted with caution. Though, several attempts were made to ensure maximal participation of all the eligible doctors, it is unclear whether volunteer bias has any effect on the findings. Therefore, future studies should aim at recruiting representative sample of doctors across the various municipalities in the province and across multiple provinces in the country.

## Conclusion

Young and early career medical doctors are less likely to be retained in the rural health facilities. These doctors prioritise career growth and development as the main determinants for retention whilst older and more experienced doctors rated improved living conditions as a major priority for retention in the rural health facilities. Surprisingly, financial incentives are no longer considered a priority as a retention strategy among the study sample. Therefore, the district health authorities should provide training support and career development of doctors. In addition, renovation and refurbishment of existing accommodation in the various district hospitals should be undertaken as a retention strategy in the rural health facilities.

## Data Availability

The minimal dataset is available at Zenodo via 10.5281/zenodo.19635202

## Acknowledgements

The author would like to acknowledge the contributions of the participants in the study along with the guidance received from their supervisors, and the District Clinical Specialist Teams of the Amathole and Buffalo City Metropolitan Health Districts.

---

**Supplementary 1:**
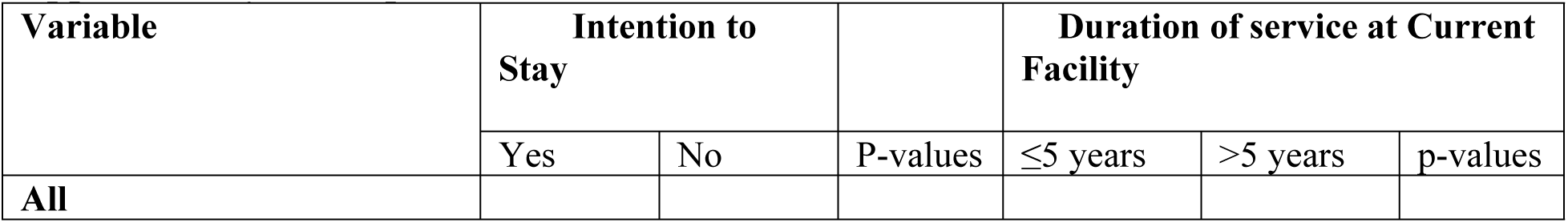
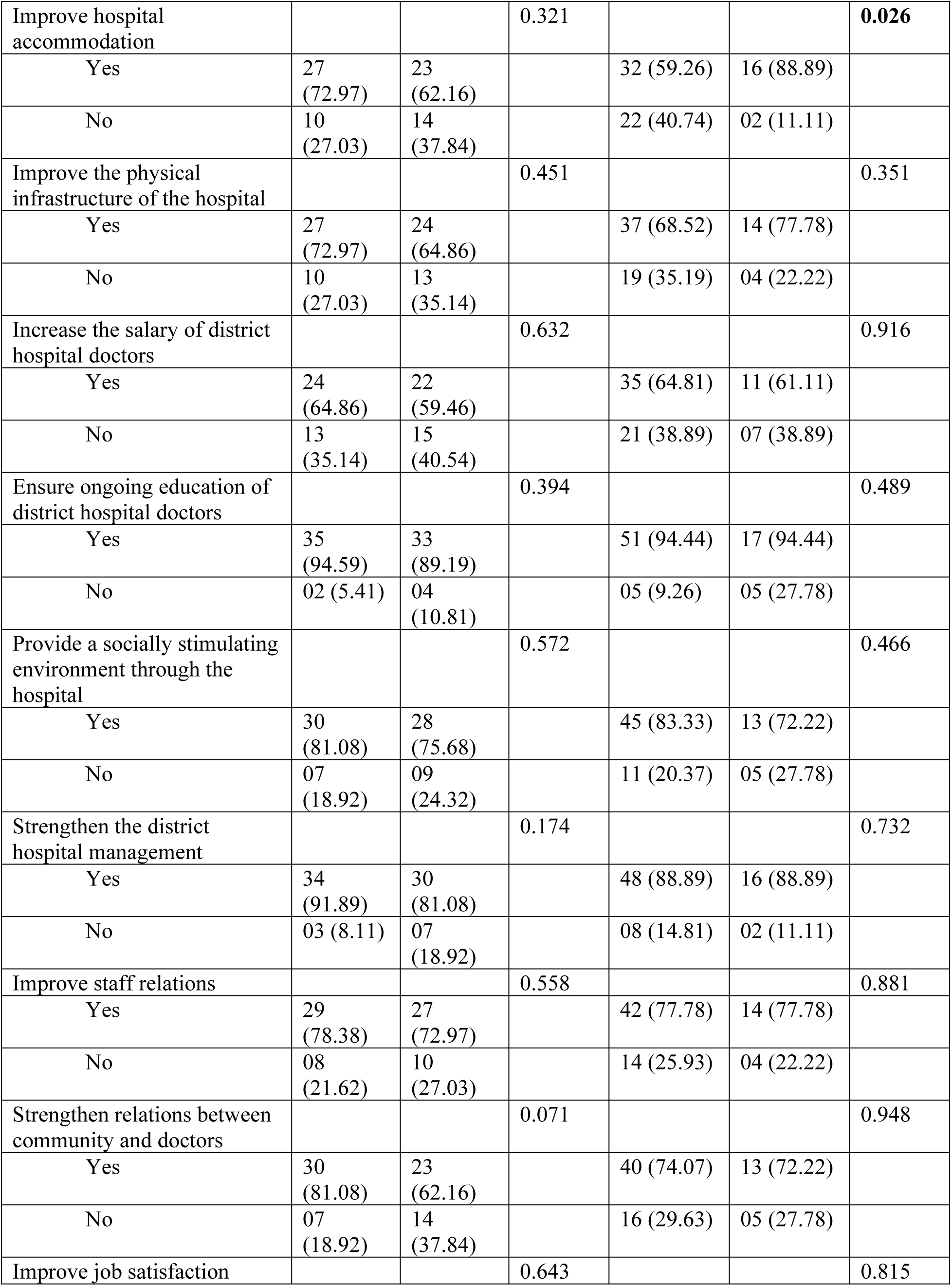
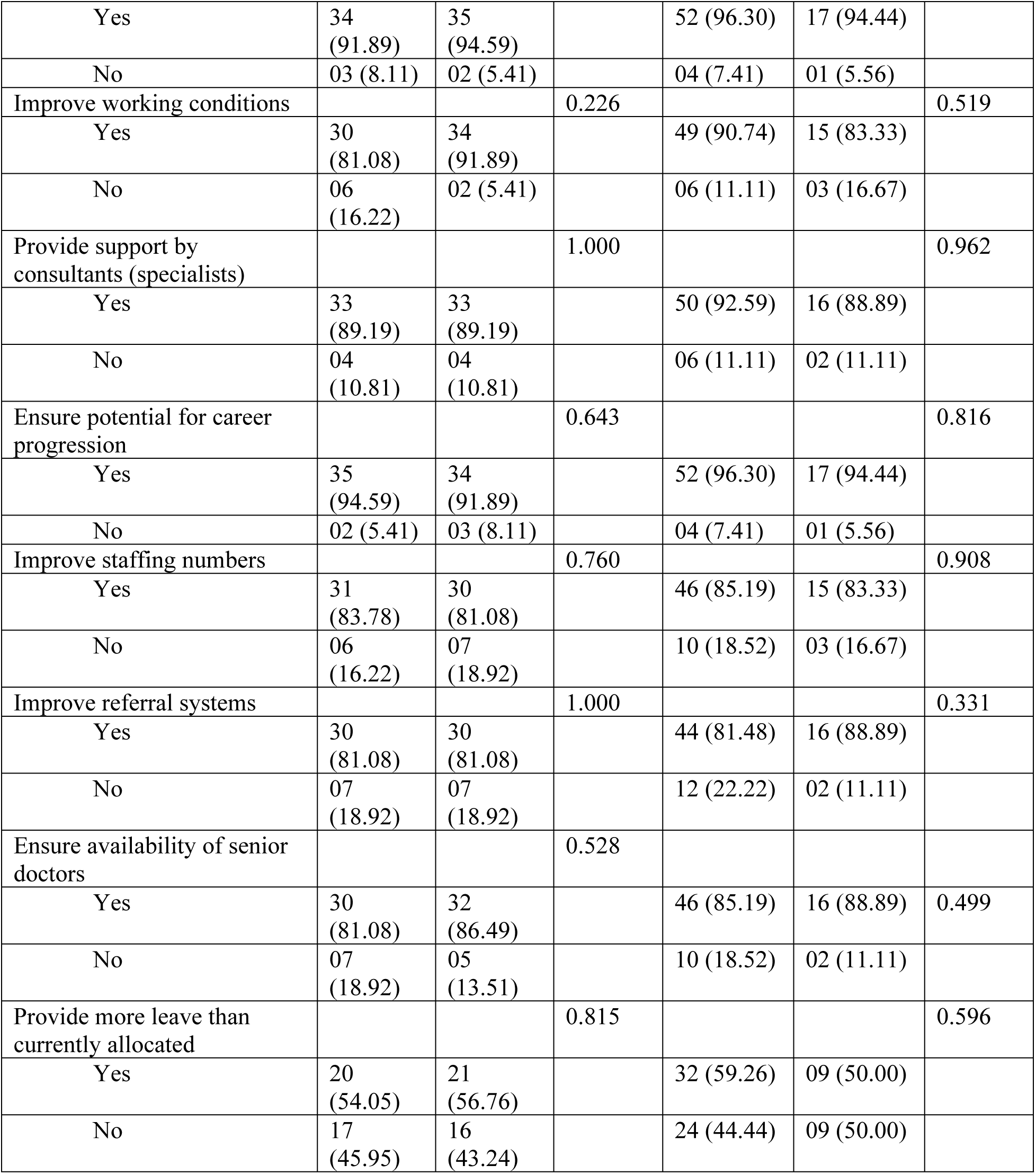
Hospital Factors.

